# Reduced Risk of Cardiovascular Diseases after Bariatric Surgery Based on the New PREVENT Equations

**DOI:** 10.1101/2024.08.05.24311527

**Authors:** Lei Wang, Xinmeng Zhang, You Chen, Charles R. Flynn, Wayne J. English, Jason M. Samuels, Brandon Williams, Matthew Spann, Vance L. Albaugh, Xiao-Ou Shu, Danxia Yu

## Abstract

**Background:** We applied the novel Predicting Risk of Cardiovascular Disease EVENTs (PREVENT) equations to evaluate cardiovascular-kidney-metabolic (CKM) health and estimated CVD risk, including heart failure (HF), after bariatric surgery.

**Methods:** Among 7804 patients (20–79 years) undergoing bariatric surgery at Vanderbilt University Medical Center during 1999–2022, CVD risk factors at pre-surgery, 1-year, and 2-year post-surgery were extracted from electronic health records. The 10- and 30-year risks of total CVD, atherosclerotic CVD (ASCVD), coronary heart disease (CHD), stroke, and HF were estimated for patients without a history of CVD or its subtypes at each time point, using the social deprivation index-enhanced PREVENT equations. Paired t-tests or McNemar tests were used to compare pre- with post-surgery CKM health and CVD risk. Two-sample t-tests were used to compare CVD risk reduction between patient subgroups defined by age, sex, race, operation type, weight loss, and history of diabetes, hypertension, and dyslipidemia.

**Results:** CKM health was significantly improved after surgery with lower systolic blood pressure, non-high-density-lipoprotein cholesterol (non-HDL), and diabetes prevalence, but higher HDL and estimated glomerular filtration rate (eGFR). The 10-year total CVD risk decreased from 6.51% at pre-surgery to 4.81% and 5.08% at 1- and 2-year post-surgery (relative reduction: 25.9% and 16.8%), respectively. Significant risk reductions were seen for all CVD subtypes (i.e., ASCVD, CHD, stroke, and HF), with the largest reduction for HF (relative reduction: 55.7% and 44.8% at 1- and 2-year post-surgery, respectively). Younger age, White race, >30% weight loss, diabetes history, and no dyslipidemia history were associated with greater HF risk reductions. Similar results were found for the 30-year risk estimates.

**Conclusions:** Bariatric surgery significantly improves CKM health and reduces estimated CVD risk, particularly HF, by 45-56% within 1-2 years post-surgery. HF risk reduction may vary by patient’s demographics, weight loss, and disease history, which warrants further research.

## Introduction

Cardiovascular diseases (CVD), including mainly coronary heart disease (CHD), stroke, and heart failure (HF), are leading causes of death, accounting for one in every five deaths in the United States (US) and one in every three deaths globally in recent years.^1–3^ Obesity, diabetes, hypertension, dyslipidemia, and chronic kidney disease (CKD) are established risk factors for CVD.^4^ Bariatric surgery, the most effective and durable treatment for obesity and its related comorbidities, offers a long-lasting strategy for mitigating CVD risk and risk factors. Bariatric surgery leads to a 30% weight loss while reducing hemoglobin A1c (HbA1c) by 0.3% to 2.7%, systolic blood pressure (SBP) by 7.2 to 15.1 mmHg, and total cholesterol (TG) by 21.6 to 66.7 mg/dL and increasing estimated glomerular filtration rate (eGFR) by an average of 14.27 ml/min/1.73 m^2^ at 1-year post-surgery.^5–10^ In addition, bariatric surgery achieves remission rates ranging from 33% to 90% for diabetes, 43% to 83% for hypertension, 16% to 80% for dyslipidemia at 1-year post-surgery, and 77% to 83% for albuminuria at 6-12 months.^11–15^

Regarding the risk of future CVD events, we recently reported a ∼34% reduction in 10-year atherosclerotic CVD (ASCVD) risk, in line with a 2020 review noting a 19% to 54% relative reduction in 10-year CVD risk one year after bariatric surgery.^9,16^ All previous studies used well-known CVD risk models such as the Framingham Risk Score for 10-year CHD risk and the American College of Cardiology/American Heart Association (ACC/AHA) pooled cohort equations for 10-year ASCVD risk. However, previous models have not accounted for the additional risk posed by reduced renal function or socioeconomic disadvantage nor predicted HF risk. Decreased eGFR, increased albuminuria, and living in an area with socioeconomic deprivation have been increasingly recognized for contributing to CVD incidence and mortality.^17–21^ In response to this gap, a new CVD risk assessment tool, termed the Predicting Risk of Cardiovascular Disease EVENTs (PREVENT) equations, was introduced by the AHA in 2023, aiming to assess total CVD risk more accurately, precisely, and equitably across diverse populations.^22^ This new tool incorporates traditional risk factors and eGFR in the base model, offers additional predictors of renal (urine albumin-to-creatinine ratio), metabolic (HbA1c), and social (social deprivation index [SDI]) risk in the add-on models for enhancement, and captures total CVD events, including HF, which was not included in previous models.^22^

Herein, by leveraging longitudinal data from the Medical and Surgical Weight Loss Clinic at Vanderbilt University Medical Center (VUMC) and Electronic Health Records (EHR) from all VUMC clinics, we evaluated post-bariatric surgery CKM health improvement and CVD risk reduction, including total CVD, ASCVD, CHD, stroke, and HF, using the novel SDI-enhanced PREVENT equations in a cohort of >7800 patients. We also assessed the potential variations in CVD risk reduction among patients with different demographics (i.e., age, sex, and race), operation types (Roux-en-Y gastric bypass [RYGB] vs. sleeve gastrectomy [SG]), weight loss amounts, and comorbidities (i.e., diabetes, hypertension, and dyslipidemia), to gain further knowledge to guide personalized management strategies and optimize post-operative care.

## Methods

### Study population and data extraction

This study was conducted in the bariatric surgery cohort at VUMC, consisting of 7804 patients who were 20 to 79 years old and underwent their first bariatric surgery between January 1999 and July 2022.^9,23^ Pre- and post-surgery data over 1 to 120 months at 3-month intervals were collected in the Vanderbilt Metabolic and Bariatric Surgery Quality, Efficacy, and Safety (QES) database, including demographics (e.g., age, sex, and race), surgery information (e.g., surgery date, surgeon, and operation type), disease history (e.g., diabetes, hypertension, and dyslipidemia), and clinical outcomes (e.g., body weight, body mass index [BMI], and HbA1c). We linked bariatric QES with VUMC EHR and extracted data up to 10 years before and after surgery, to minimize missing data and obtain additional information on zip code for deriving SDI, smoking status, blood pressure, blood lipids, blood glucose, eGFR, disease diagnosis, and medication records. The study was approved by the VUMC Institutional Review Board, with written informed consent waived for participation because it is a retrospective study based on existing data with minimal risk to the participants.

### CVD risk calculation

Based on the new sex-specific, race-free, SDI-enhanced PREVENT equations, we estimated the 10- and 30-year CVD risk at pre-surgery, 1-year, and 2-year post-surgery, respectively, among patients without a diagnosis of CVD or its subtypes at each time point. The components for calculating the risk of total CVD, ASCVD, CHD, and stroke included age, sex, SDI, smoking status, SBP, high-density-lipoprotein cholesterol (HDL), non-HDL, eGFR, diabetes status, anti-hypertensive use, and statin use; while age, sex, SDI, smoking status, BMI, SBP, eGFR, diabetes status, and anti-hypertensive use were utilized for calculating the risk of HF. We defined 6-month time windows to fully capture the values of those components at pre-surgery (i.e., 6 months to 1 week before surgery), 1-year post-surgery (i.e., 9 to 15 months after surgery), and 2-year post-surgery (i.e., 21 to 27 months after surgery). The median value of all measurements during the 6-month time window was used for calculation, after excluding outliers that fell below Q1−1.5 interquartile range (IQR) or above Q3+1.5 IQR. Due to the requirement for smoking cessation prior to bariatric surgery, we extended the pre-surgery time window by 6 months to assess smoking status (i.e., 1 year to 1 week before surgery). In addition, anti-hypertensive use or statin use was defined as any recorded use of any anti-hypertensive drug or statin during the 6-month time window. Diabetes status was defined if any of the following indications was found: International Classification of Disease [ICD]-9: 250/ ICD-10-CM: E10-E11, using diabetes treatments, random glucose ≥200 mg/dL, or HbA1c ≥ 6.5%.

### Statistical analysis

Participants with pre- and post-surgery measures for estimating CVD risk were included. Components for calculating ASCVD risk and HF risk at pre- and post-surgery were summarized as mean (SD) for continuous variables and number (percentage) for categorical variables and compared using paired t-tests or McNemar tests. Pre- vs. 1-year and 2-year post-surgery CVD risk were compared by paired t-tests; results were presented as absolute reduction (95% confidence interval [CI] and relative reduction [95% CI]). Subgroup analyses for assessing the relative reduction of CVD risk were conducted by age (≤45 vs. > 45 years), sex (women vs. men), race (White vs. Black), operation (RYGB vs. SG), weight loss at 1- or 2-year post-surgery (≤30% vs. >30%), and pre-surgery history of diabetes (no vs. yes), hypertension (no vs. yes), and dyslipidemia (no vs. yes); two-sample t-tests were used for assessing heterogeneity. Two-sided *P* < 0.05 was considered statistically significant. All analyses were performed in SAS (version 9.4; SAS Institute, Cary, NC, USA).

## Results

### The PREVENT equations

**Table 1** presents the pre- and post-surgery values of the nine components included in the HF risk model and eleven components in the ASCVD risk model. A total of 2,910 patients were included to compare the HF risk at 1-year post-surgery vs. pre-surgery. Of these patients (82% women), the mean age was 46.0 (SD: 10.8) years. Compared with pre-surgery, patients had significant improvements in BMI (47.3 vs. 32.4 kg/m^2^), SBP (134.3 vs. 123.6 mmHg), and eGFR (93.2 vs. 100.2 mL/min/1.73 m^2^) at 1-year post-surgery; the prevalence of diabetes (43.4% vs. 35.1%) and anti-hypertensive drug use (62.6% vs. 49.4%) also decreased at 1-year post-surgery (all *P* <0.0001). Sustained CKM improvements were observed at 2-year post-surgery among 1,478 patients. For ASCVD risk, 252 and 130 patients were included for 1- and 2-year post-surgery vs. pre-surgery comparisons. In addition to improvements in SBP and eGFR, patients showed significant improvements in blood lipids, including both non-HDL (3.44 vs. 2.82 mmol/L, *P*<0.0001) and HDL (1.23 vs. 1.50 mmol/L, *P*<0.0001) at 1-year post-surgery, which were also observed at 2-year post-surgery.

**Table 1.** Components of SDI-enhanced PREVENT equations for predicting the 10-year HF and ASCVD risk.

| Components | HF risk model |  |  |  |  |  |
| --- | --- | --- | --- | --- | --- | --- |
|  | Pre-surgery vs. 1-year post-surgery<br>(N=2910) |  |  | Pre-surgery vs. 2-year post-surgery<br>(N=1478) |  |  |
|  |  |  | *P* value |  |  | *P* value |
| Age, years | 46.0 (10.8) | / | / | 47.0 (10.6) | / | / |
| Sex, men | 523 (18.0%) | / | / | 242 (16.4%) | / | / |
| SDI decile |  |  | / |  |  | / |
| 1 - 3 | 866 (29.8%) | / |  | 451 (30.5%) | / |  |
| 4 - 6 | 798 (27.4%) | / |  | 438 (29.6%) | / |  |
| 7 - 10 | 1008 (34.6%) | / |  | 513 (34.7%) | / |  |
| Missing | 238 (8.2%) | / |  | 76 (5.1%) | / |  |
| Current smoking | 140 (4.8%) | 23 (0.8%) | <.0001 | 60 (4.1%) | 25 (1.7%) | <.0001 |
| BMI, kg/m<sup>2</sup> | 47.3 (7.6) | 32.4 (6.3) | <.0001 | 46.8 (7.4) | 31.9 (6.4) | <.0001 |
| SBP, mmHg | 134.3 (12.4) | 123.6 (13.9) | <.0001 | 134.1 (12.5) | 124.2 (14.3) | <.0001 |
| eGFR, mL/min/1.73 m<sup>2</sup> | 93.2 (22.4) | 100.2 (22.4) | <.0001 | 93.3 (23.3) | 98.0 (22.7) | <.0001 |
| Diabetes | 1263 (43.4%) | 1021 (35.1%) | <.0001 | 687 (46.5%) | 614 (41.5%) | <.0001 |
| Anti-hypertensive use | 1821 (62.6%) | 1437 (49.4%) | <.0001 | 991 (67.1%) | 807 (54.6%) | <.0001 |
| Components | ASCVD risk model |  |  |  |  |  |
|  | Pre-surgery vs. 1-year post-surgery<br>(N=252) |  |  | Pre-surgery vs. 2-year post-surgery<br>(N=130) |  |  |
|  |  |  | *P* value |  |  | *P* value |
| Age, years | 46.8 (9.6) | / | / | 46.5 (9.9) | / | / |
| Sex, men | 55 (21.8%) | / | / | 22 (16.9%) | / | / |
| SDI decile |  |  | / |  |  | / |
| 1 - 3 | 77 (30.6%) | / |  | 44 (33.9%) | / |  |
| 4 - 6 | 61 (24.2%) | / |  | 35 (26.9%) | / |  |
| 7 - 10 | 70 (27.8%) | / |  | 36 (27.7%) | / |  |
| Missing | 44 (17.5%) | / |  | 15 (11.5%) | / |  |
| **Current smoking** | 6 (2.4%) | 0 (0.0%) | / | 7 (5.4%) | 3 (2.3%) | 0.29 |
| **SBP, mmHg** | 133.3 (12.4) | 124.9 (14.0) | <.0001 | 133.4 (11.6) | 125.8 (14.4) | <.0001 |
| **HDL, mmol/L** | 1.23 (0.30) | 1.50 (0.34) | <.0001 | 1.27 (0.33) | 1.59 (0.40) | <.0001 |
| **Non-HDL, mmol/L** | 3.44 (0.84) | 2.82 (0.80) | <.0001 | 3.46 (0.77) | 2.91 (0.85) | <.0001 |
| **eGFR, mL/min/1.73 m<sup>2</sup>** | 93.1 (23.0) | 100.3 (23.5) | <.0001 | 92.2 (23.1) | 97.7 (23.3) | 0.002 |
| **Diabetes** | 150 (59.5%) | 125 (49.6%) | <.0001 | 72 (55.4%) | 69 (53.1%) | 0.47 |
| **Anti-hypertensive use** | 159 (63.1%) | 139 (55.2%) | 0.002 | 94 (72.3%) | 77 (59.2%) | 0.002 |
| **Statin use** | 102 (40.5%) | 84 (33.3%) | 0.0007 | 56 (43.1%) | 45 (34.6%) | 0.01 |

Abbreviations: ASCVD: atherosclerotic cardiovascular disease, BMI: body mass index, eGFR: estimated glomerular filtration rate, HDL: high-density lipoprotein, HF: heart failure, PREVENT: Predicting Risk of Cardiovascular Disease EVENTS, SBP: systolic blood pressure, SDI: social deprivation index.
\* Data was presented as mean (SD) or Number (%).

### CVD risk reduction after surgery

Substantial reductions in estimated 10-year and 30-year CVD risk were observed after surgery (**Table 2**). Specifically, the estimated 10-year total CVD risk decreased from 6.51% at pre-surgery to 4.81% at 1-year post-surgery, corresponding to a relative reduction of 25.90% (95% CI: 19.79%, 32.02%). Regarding CVD subtypes, relative reductions of 28.36% (95% CI: 22.43%, 34.30%), 32.85% (26.99%, 38.72%), 20.74% (15.47%, 26.01%), and 55.68% (54.36%, 57.00%) were observed for ASCVD, CHD, stroke, and HF risk, respectively. Sustainable risk reductions, ranging from 12.09% to 44.76%, were observed at 2-year post-surgery, similarly, with the least reduction observed for stroke and the greatest reduction for HF.

**Table 2.**
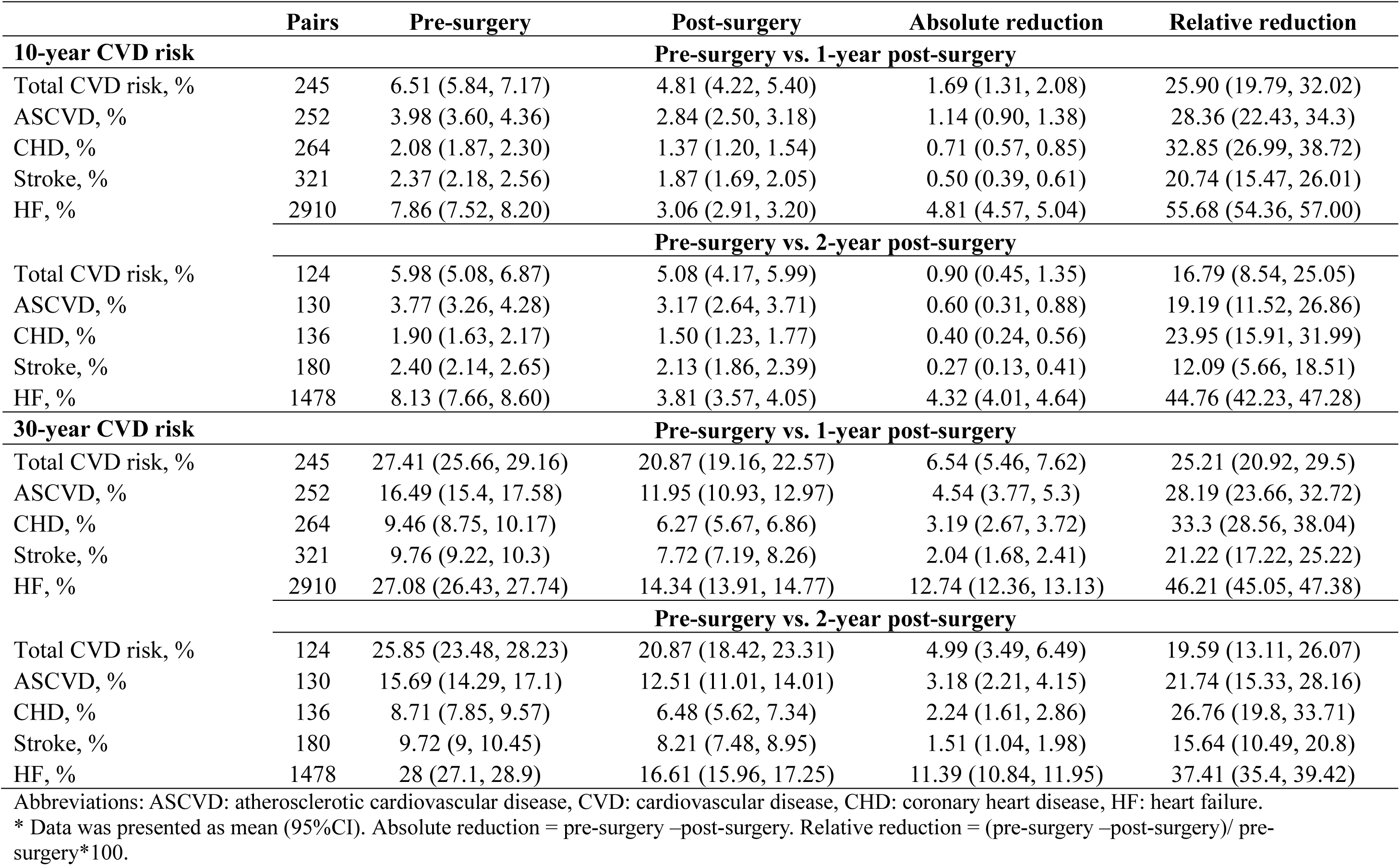
Comparison of CVD risk between pre- and post-surgery.

|  | Pairs | Pre-surgery | Post-surgery | Absolute reduction | Relative reduction |
| --- | --- | --- | --- | --- | --- |
| **10-year CVD risk** |  | **Pre-surgery vs. 1-year post-surgery** |  |  |  |
| Total CVD risk, % | 245 | 6.51 (5.84, 7.17) | 4.81 (4.22, 5.40) | 1.69 (1.31, 2.08) | 25.90 (19.79, 32.02) |
| ASCVD, % | 252 | 3.98 (3.60, 4.36) | 2.84 (2.50, 3.18) | 1.14 (0.90, 1.38) | 28.36 (22.43, 34.3) |
| CHD, % | 264 | 2.08 (1.87, 2.30) | 1.37 (1.20, 1.54) | 0.71 (0.57, 0.85) | 32.85 (26.99, 38.72) |
| Stroke, % | 321 | 2.37 (2.18, 2.56) | 1.87 (1.69, 2.05) | 0.50 (0.39, 0.61) | 20.74 (15.47, 26.01) |
| HF, % | 2910 | 7.86 (7.52, 8.20) | 3.06 (2.91, 3.20) | 4.81 (4.57, 5.04) | 55.68 (54.36, 57.00) |
|  |  | **Pre-surgery vs. 2-year post-surgery** |  |  |  |
| Total CVD risk, % | 124 | 5.98 (5.08, 6.87) | 5.08 (4.17, 5.99) | 0.90 (0.45, 1.35) | 16.79 (8.54, 25.05) |
| ASCVD, % | 130 | 3.77 (3.26, 4.28) | 3.17 (2.64, 3.71) | 0.60 (0.31, 0.88) | 19.19 (11.52, 26.86) |
| CHD, % | 136 | 1.90 (1.63, 2.17) | 1.50 (1.23, 1.77) | 0.40 (0.24, 0.56) | 23.95 (15.91, 31.99) |
| Stroke, % | 180 | 2.40 (2.14, 2.65) | 2.13 (1.86, 2.39) | 0.27 (0.13, 0.41) | 12.09 (5.66, 18.51) |
| HF, % | 1478 | 8.13 (7.66, 8.60) | 3.81 (3.57, 4.05) | 4.32 (4.01, 4.64) | 44.76 (42.23, 47.28) |
| **30-year CVD risk** |  | **Pre-surgery vs. 1-year post-surgery** |  |  |  |
| Total CVD risk, % | 245 | 27.41 (25.66, 29.16) | 20.87 (19.16, 22.57) | 6.54 (5.46, 7.62) | 25.21 (20.92, 29.5) |
| ASCVD, % | 252 | 16.49 (15.4, 17.58) | 11.95 (10.93, 12.97) | 4.54 (3.77, 5.3) | 28.19 (23.66, 32.72) |
| CHD, % | 264 | 9.46 (8.75, 10.17) | 6.27 (5.67, 6.86) | 3.19 (2.67, 3.72) | 33.3 (28.56, 38.04) |
| Stroke, % | 321 | 9.76 (9.22, 10.3) | 7.72 (7.19, 8.26) | 2.04 (1.68, 2.41) | 21.22 (17.22, 25.22) |
| HF, % | 2910 | 27.08 (26.43, 27.74) | 14.34 (13.91, 14.77) | 12.74 (12.36, 13.13) | 46.21 (45.05, 47.38) |
|  |  | **Pre-surgery vs. 2-year post-surgery** |  |  |  |
| Total CVD risk, % | 124 | 25.85 (23.48, 28.23) | 20.87 (18.42, 23.31) | 4.99 (3.49, 6.49) | 19.59 (13.11, 26.07) |
| ASCVD, % | 130 | 15.69 (14.29, 17.1) | 12.51 (11.01, 14.01) | 3.18 (2.21, 4.15) | 21.74 (15.33, 28.16) |
| CHD, % | 136 | 8.71 (7.85, 9.57) | 6.48 (5.62, 7.34) | 2.24 (1.61, 2.86) | 26.76 (19.8, 33.71) |
| Stroke, % | 180 | 9.72 (9, 10.45) | 8.21 (7.48, 8.95) | 1.51 (1.04, 1.98) | 15.64 (10.49, 20.8) |
| HF, % | 1478 | 28 (27.1, 28.9) | 16.61 (15.96, 17.25) | 11.39 (10.84, 11.95) | 37.41 (35.4, 39.42) |

Abbreviations: ASCVD: atherosclerotic cardiovascular disease, CVD: cardiovascular disease, CHD: coronary heart disease, HF: heart failure.
\* Data was presented as mean (95%CI). Absolute reduction = pre-surgery –post-surgery. Relative reduction = (pre-surgery –post-surgery)/ pre-surgery\*100.

The estimated 30-year total CVD risk was 27.41% pre-surgery, which reduced to 20.87% at both 1- and 2-year post-surgery. The relative reductions of the 30-year total CVD risk and its subtypes ranged from 21.22% to 46.21% at 1-year and from 15.64% to 37.41% at 2-year post-surgery, with the least reduction observed for stroke and the greatest reduction for HF.

### CVD risk reduction among patient subgroups

Similar reductions in the estimated 10-year risks of total CVD, ASCVD, CHD, and stroke were observed at 1-year post-surgery among male and female patients, Black and White patients, RYGB and VSG treated patients, and those with/without a history of diabetes, hypertension, or dyslipidemia (**Figure 1**), while more evident in patients ≤ 45 years old (37.25% reduction in total CVD risk) or those with >30% weight loss (30.92% reduction in total CVD risk) than their counterparts (all *P* for heterogeneity ≤0.05; **Figure 1** and **Supplementary Table 1**). Results remained similar after mutually adjusting for age, sex, race, operation type, surgery year, weight loss, cardiometabolic disease history, and pre-surgery CVD risk (**Supplementary Table 1**). Of note, 10-year HF risk reduction was greater among younger patients (58.05%), White patients (56.73%), and those with >30% weight loss (60.76%), had a history of diabetes (63.16%) or hypertension (56.68%), or had no history of dyslipidemia (57.05%) than their counterparts with or without mutual adjustment (all *P* for heterogeneity <0.05); nevertheless, HF risk reduction was significant and substantial across all patient subgroups (**Figure 1** and **Supplementary Table 1**).

**Figure 1.**
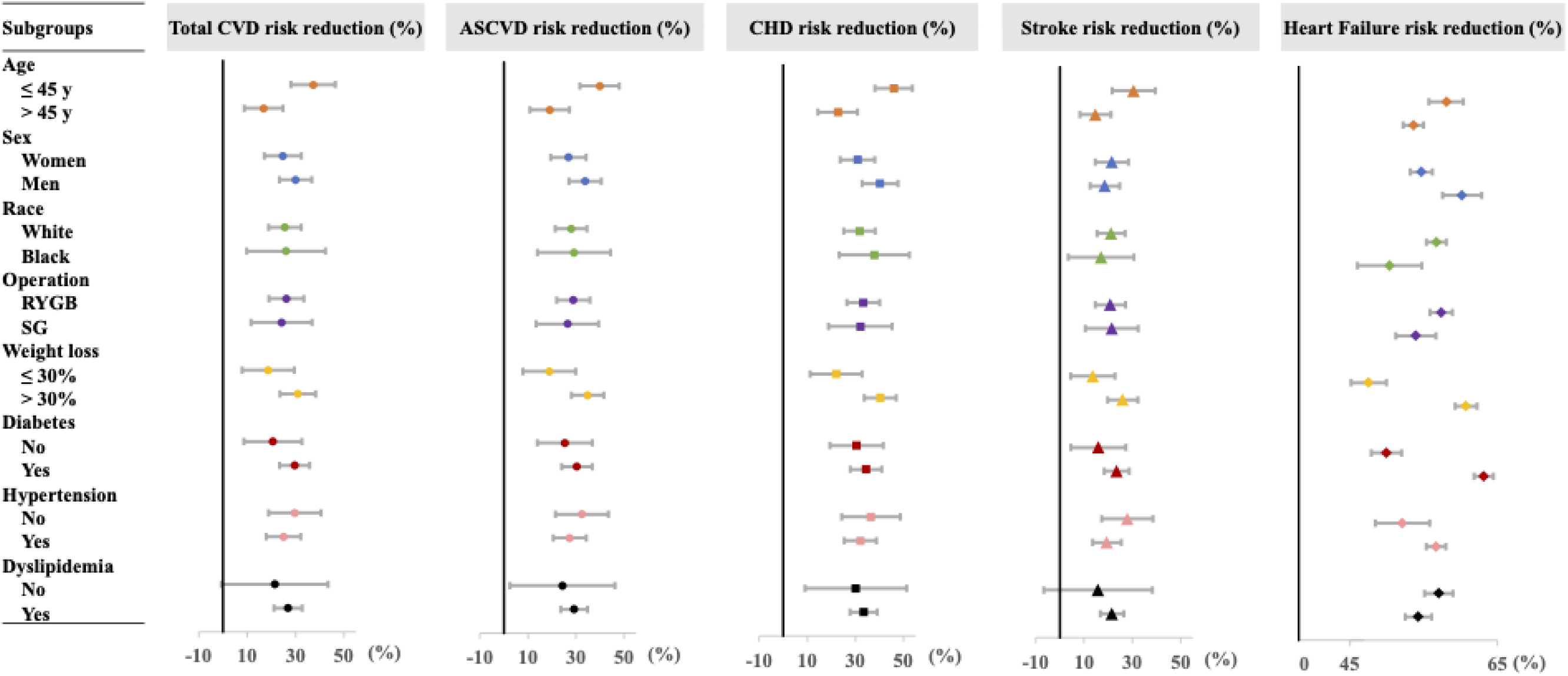
The relative reduction of 10-year CVD risk in subgroups at 1-year post-surgery. Abbreviations: ASCVD: atherosclerotic cardiovascular disease, CVD: cardiovascular disease, CHD: coronary heart disease, RYGB: Roux-en-Y gastric bypass, SG: sleeve gastrectomy.

At 2-year post-surgery, reductions in the 10-year risk of total CVD, ASCVD, CHD, and stroke were all similar between patient subgroups; however, for 10-year HF risk, patients who were younger, White, achieved >30% weight loss, had a history of diabetes, or had no history of dyslipidemia remained showed greater reductions than their counterparts after mutual adjustment (all *P* for heterogeneity <0.05, **Figure 2** and **Supplementary Table 2**).

**Figure 2.**
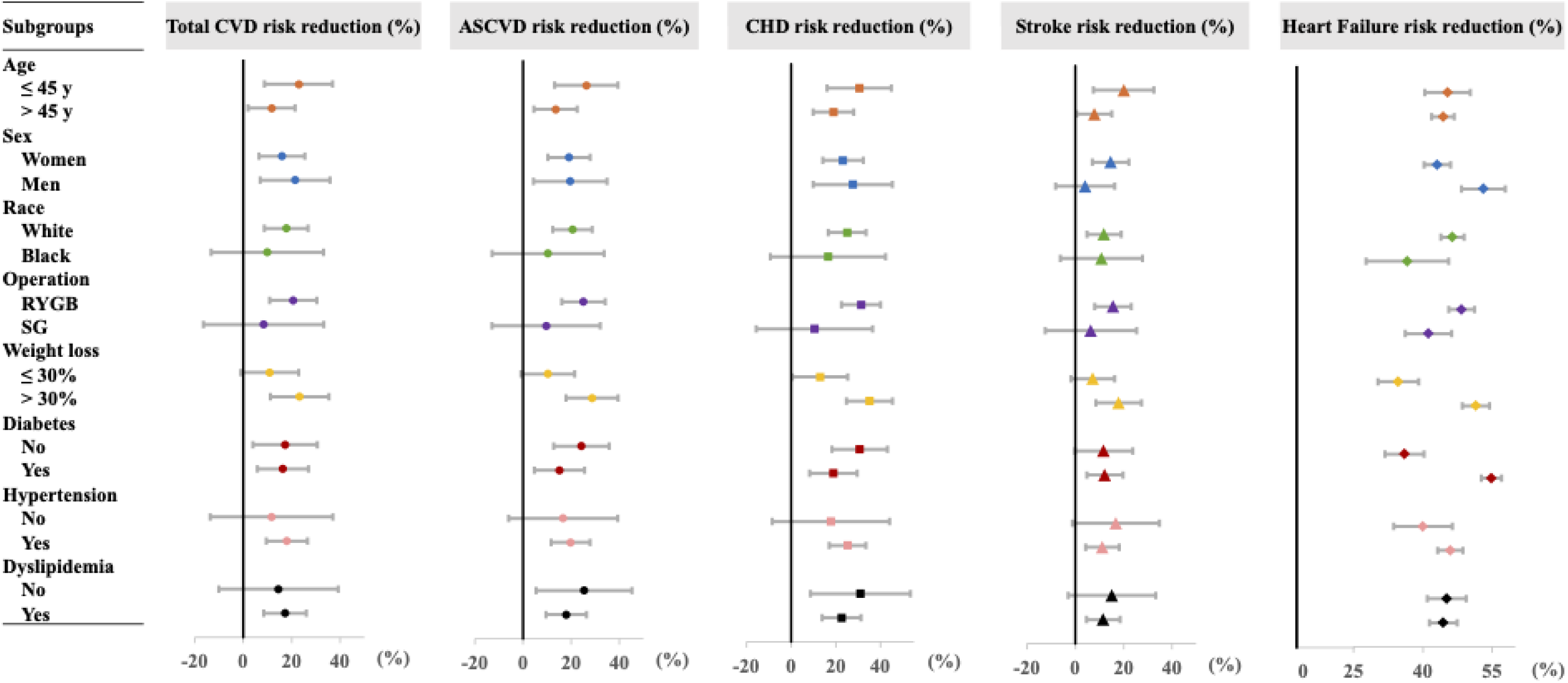
The relative reduction of 10-year CVD risk in subgroups at 2-year post-surgery. Abbreviations: ASCVD: atherosclerotic cardiovascular disease, CVD: cardiovascular disease, CHD: coronary heart disease, RYGB: Roux-en-Y gastric bypass, SG: sleeve gastrectomy.

Similar patterns were observed for the estimated 30-year CVD risk (**Supplementary Figures 1 & 2** and **Supplementary Tables 3 & 4**).

## Discussion

In this single-center EHR-based, longitudinal cohort of bariatric surgery patients, we observed significant improvements in CKM health after surgery, including reduced BMI, SBP, non-HDL, diabetes prevalence, and use of anti-hypertensive drugs and statins, as well as increased HDL and eGFR. The estimated 10-year risks of total CVD, ASCVD, CHD, and stroke decreased by 21% to 33% and 12% to 24% at 1-year and 2-year post-surgery, and remarkably, 56% and 45% reductions were observed for estimated 10-year HF risk at 1-year and 2-year post-surgery, respectively. Furthermore, we found that younger age, White race, >30% weight loss, diabetes history, and no dyslipidemia history were associated with greater reductions in HF risk after bariatric surgery.

Although the significant CVD benefits of bariatric surgery have been well recognized, to our knowledge, this is the first study applying the new sex-specific, race-free PREVENT equations, which include SDI and eGFR as predictors to capture patients’ socioeconomic status and overall CKM health and predict 10-year and 30-year HF risk. Our findings on CKM health improvements were consistent with the literature and results reported in our recent publication, which showed significant reductions in blood pressure, blood lipids, glucose, and HbA1c, as well as 32% to 50% remissions rates of diabetes, hypertension, and dyslipidemia 1 year after bariatric surgery.^11,24,9^ In addition, we observed 28% and 19% reductions in 10-year ASCVD risk at 1- and 2-year post-surgery, which is in line with previous reports, ranging from 19% to 54%,^16^ although the reductions appear to be smaller than our previous results based on the ACC/AHA equations (34% and 23%).^9^ This discrepancy could be explained by differences in those CVD risk prediction models. The new PREVENT equations were derived among >6 million people aged 30-79 years from 46 cohorts,^22^ while the ACC/AHA equations were race-specific risk prediction models developed in ∼25,000 non-Hispanic White and Black people aged 40-79 years from only 5 cohorts.^25^ In addition, the PREVENT equations newly included eGFR as a predictor, which has been associated with increased risk of ASCVD, independent of traditional CVD risk factors.^17^

We observed the greatest CVD benefit in HF prevention, with a 45-56% risk reduction within 1-2 years after surgery. Several studies have suggested reduced HF risk after bariatric surgery,^26–30^ including a 2018 meta-analysis reporting a hazard ratio of 0.44 (95% CI: 0.36-0.55) for incident HF following bariatric surgery compared with non-surgical controls.^30^ The dramatic reduction of HF risk is likely driven by the decrease in BMI (47.3 kg/m^2^ at pre-surgery to 32.4 kg/m^2^ at 1-year post-surgery in our study sample), a strong predictor in the PREVENT equations for HF risk. In a pooled analysis of 10 prospective cohorts in the US, obesity was significantly associated with increased CVD morbidity and mortality; of note, BMI showed the strongest association with incident HF, around a 5-fold increase compared with other CVD subtypes.^31^

We further observed that HF risk reduction was greater among younger or White patients than among older or Black patients. Previous studies have linked younger age with more pronounced cardiometabolic improvements after surgery, consistent with our findings.^9,32,33^ However, evidence regarding effect modification by race has been inconsistent. Our previous results showed a greater reduction in ASCVD risk in White patients than in Black patients, while another study reported the opposite.^9,34^ Over 30% weight loss was also associated with a greater HF risk reduction in our study, which is expected given the central role of obesity in CKM health and CVD etiology.^35^ In the Swedish Obese Subjects cohort study, patients in the highest quartile of weight loss after 1 year (mean: 41 kg) showed the greatest reduction of incident HF (∼40%).^29^ We also found that patients with a history of diabetes or no history of dyslipidemia showed greater HF risk reductions than their counterparts, which seems contradictory. Previous research has generally suggested more cardiometabolic improvements and ASCVD risk reduction among patients without a history of metabolic diseases.^9,33,36,37^ Currently, few HF risk prediction models have been developed and externally validated,^38^ and few studies have applied HF risk models among bariatric surgery patients. Thus, although substantial HF risk reductions were observed in all patient subgroups in our study (48-63% 1-year post-surgery), future studies are needed to clarify the potential heterogenetic effects of bariatric surgery on HF risk by race and comorbidities.

This study has considerable strengths. First, we applied the novel PREVENT equations, which newly added eGFR as a predictor, to fully capture the CKM benefits of bariatric surgery. Second, we used the SDI-enhanced PREVENT equations to capture social determinants of health. Third, we had a large sample size (n = ∼3000 for estimating the HF risk) and noted sustained CKM health improvements and HF risk reduction 2 years following bariatric surgery. Fourth, we utilized EHR data from all VUMC clinics to improve data completeness and quality. There are also several limitations. First, the sample sizes for estimating the total CVD and ASCVD risk were much smaller than the ones for HF risk. Total CVD and ASCVD risk estimation required blood lipid data, which were not routinely measured for patients with obesity. Small sample sizes may lead to insufficient statistical power to detect differences between patient subgroups, although we observed significant risk reductions among total patients at both 1-year and 2-year post-surgery. Second, given the observational and retrospective nature, our results may be affected by confounding and selection biases. Future studies with large sample sizes, long-term follow-ups, and comprehensive measurements are needed to evaluate the sustainability of CKM and CVD benefits and the potential heterogeneity effect of bariatric surgery in patient subgroups.

In conclusion, our study demonstrated that bariatric surgery significantly improved CKM health and reduced the risk of total CVD and its subtypes, particularly HF, with an estimated 45-56% risk reduction at 1-2 years after surgery. HF risk reduction may vary based on patients’ demographics and disease history, which warrants further research.

## Data Availability

Restrictions apply to the availability of some or all data generated or analyzed during this study to preserve patient confidentiality or because they were used under license. The corresponding author will on request detail the restrictions and any conditions under which access to some data may be provided.

## Acknowledgments

LW and DY designed the study. XZ, YC, CRF, VA, WJE, BW, MS, and JS collected the data. LW cleaned and analyzed data. LW drafted the manuscript. All authors contributed to reviewing and editing the paper and approved the final version of the manuscript. DY is the guarantor of the work and takes responsibility for its integrity and accuracy.

## Reference

1. Martin SS, Aday AW, Almarzooq ZI, Anderson CAM, Arora P, Avery CL, Baker-Smith CM, Barone Gibbs B, Beaton AZ, Boehme AK, et al. 2024 Heart Disease and Stroke Statistics: A Report of US and Global Data From the American Heart Association. Circulation. 2024;149(8). doi:10.1161/CIR.0000000000001209

2. National Center for Health Statistics. Multiple Cause of Death 2018–2022 on CDC WONDER Database. Accessed July 9, 2024. https://wonder.cdc.gov/mcd.html

3. Lindstrom M, DeCleene N, Dorsey H, Fuster V, Johnson CO, LeGrand KE, Mensah GA, Razo C, Stark B, Varieur Turco J, et al. Global Burden of Cardiovascular Diseases and Risks Collaboration, 1990-2021. Journal of the American College of Cardiology. 2022;80(25):2372–2425. doi:10.1016/j.jacc.2022.11.001

4. Bays HE, Taub PR, Epstein E, Michos ED, Ferraro RA, Bailey AL, Kelli HM, Ferdinand KC, Echols MR, Weintraub H, et al. Ten things to know about ten cardiovascular disease risk factors. American Journal of Preventive Cardiology. 2021;5:100149. doi:10.1016/j.ajpc.2021.100149

5. Van Rijswijk AS, Van Olst N, Schats W, Van Der Peet DL, Van De Laar AW. What Is Weight Loss After Bariatric Surgery Expressed in Percentage Total Weight Loss (%TWL)? A Systematic Review. OBES SURG. 2021;31(8):3833–3847. doi:10.1007/s11695-021-05394-x

6. Barzin M, Motamedi MAK, Serahati S, Khalaj A, Arian P, Valizadeh M, Khalili D, Azizi F, Hosseinpanah F. Comparison of the Effect of Gastric Bypass and Sleeve Gastrectomy on Metabolic Syndrome and its Components in a Cohort: Tehran Obesity Treatment Study (TOTS). OBES SURG. 2017;27(7):1697–1704. doi:10.1007/s11695-016-2526-0

7. Lee W, Chong K, Aung L, Chen S, Ser K, Lee Y. Metabolic Surgery for Diabetes Treatment: Sleeve Gastrectomy or Gastric Bypass? World j surg. 2017;41(1):216–223. doi:10.1007/s00268-016-3690-z

8. Benaiges D, Goday A, Ramon JM, Hernandez E, Pera M, Cano JF. Laparoscopic sleeve gastrectomy and laparoscopic gastric bypass are equally effective for reduction of cardiovascular risk in severely obese patients at one year of follow-up. Surgery for Obesity and Related Diseases. 2011;7(5):575–580. doi:10.1016/j.soard.2011.03.002

9. Wang L, O’Brien MT, Zhang X, Chen Y, English WJ, Williams B, Spann M, Albaugh V, Shu XO, Flynn CR, et al. Cardiometabolic Improvements After Metabolic Surgery and Related Presurgery Factors. Journal of the Endocrine Society. 2024;8(5):bvae027. doi:10.1210/jendso/bvae027

10. Huang H, Lu J, Dai X, Li Z, Zhu L, Zhu S, Wu L. Improvement of Renal Function After Bariatric Surgery: a Systematic Review and Meta-analysis. OBES SURG. 2021;31(10):4470–4484. doi:10.1007/s11695-021-05630-4

11. Arterburn DE, Telem DA, Kushner RF, Courcoulas AP. Benefits and Risks of Bariatric Surgery in Adults: A Review. JAMA. 2020;324(9):879–887. doi:10.1001/jama.2020.12567

12. Climent E, Goday A, Pedro-Botet J, Solà I, Oliveras A, Ramón JM, Flores-Le Roux JA, Checa MÁ, Benaiges D. Laparoscopic Roux-en-Y gastric bypass versus laparoscopic sleeve gastrectomy for 5-year hypertension remission in obese patients: a systematic review and meta-analysis. J Hypertens. 2020;38(2):185–195. doi:10.1097/HJH.0000000000002255

13. Lee Y, Doumouras AG, Yu J, Aditya I, Gmora S, Anvari M, Hong D. Laparoscopic Sleeve Gastrectomy Versus Laparoscopic Roux-en-Y Gastric Bypass: A Systematic Review and Meta-analysis of Weight Loss, Comorbidities, and Biochemical Outcomes From Randomized Controlled Trials. Ann Surg. 2021;273(1):66–74. doi:10.1097/SLA.0000000000003671

14. Cohen RV, Pereira TV, Aboud CM, Petry TBZ, Lopes Correa JL, Schiavon CA, Pompílio CE, Pechy FNQ, Da Costa Silva ACC, De Melo FLG, et al. Effect of Gastric Bypass vs Best Medical Treatment on Early-Stage Chronic Kidney Disease in Patients With Type 2 Diabetes and Obesity: A Randomized Clinical Trial. JAMA Surg. 2020;155(8):e200420. doi:10.1001/jamasurg.2020.0420

15. Fathy E, Aisha HAA, Abosayed AK, ElAnsary AMSEO, Al Aziz AA. Effect of Bariatric Surgery on Albuminuria in Non-Diabetic Non-Hypertensive Patients with Severe Obesity: a Short-Term Outcome. OBES SURG. 2022;32(7):2397–2402. doi:10.1007/s11695-022-06091-z

16. English WJ, Spann MD, Aher CV, Williams DB. Cardiovascular risk reduction following metabolic and bariatric surgery. Ann Transl Med. 2020;8(S1):S12–S12. doi:10.21037/atm.2020.01.88

17. Matsushita K, Jassal SK, Sang Y, Ballew SH, Grams ME, Surapaneni A, Arnlov J, Bansal N, Bozic M, Brenner H, et al. Incorporating kidney disease measures into cardiovascular risk prediction: Development and validation in 9 million adults from 72 datasets. EClinicalMedicine. 2020;27:100552. doi:10.1016/j.eclinm.2020.100552

18. Matsushita K, Coresh J, Sang Y, Chalmers J, Fox C, Guallar E, Jafar T, Jassal SK, Landman GWD, Muntner P, et al. Estimated glomerular filtration rate and albuminuria for prediction of cardiovascular outcomes: a collaborative meta-analysis of individual participant data. The Lancet Diabetes & Endocrinology. 2015;3(7):514–525. doi:10.1016/S2213-8587(15)00040-6

19. Chronic Kidney Disease Prognosis Consortium, Matsushita K, van der Velde M, Astor BC, Woodward M, Levey AS, de Jong PE, Coresh J, Gansevoort RT. Association of estimated glomerular filtration rate and albuminuria with all-cause and cardiovascular mortality in general population cohorts: a collaborative meta-analysis. Lancet. 2010;375(9731):2073–2081. doi:10.1016/S0140-6736(10)60674-5

20. Jankowski J, Floege J, Fliser D, Böhm M, Marx N. Cardiovascular Disease in Chronic Kidney Disease: Pathophysiological Insights and Therapeutic Options. Circulation. 2021;143(11):1157–1172. doi:10.1161/CIRCULATIONAHA.120.050686

21. Deng K, Xu M, Sahinoz M, Cai Q, Shrubsole MJ, Lipworth L, Gupta DK, Dixon DD, Zheng W, Shah R, et al. Associations of neighborhood sociodemographic environment with mortality and circulating metabolites among low-income black and white adults living in the southeastern United States. BMC Med. 2024;22(1):249. doi:10.1186/s12916-024-03452-6

22. Khan SS, Matsushita K, Sang Y, Ballew SH, Grams ME, Surapaneni A, Blaha MJ, Carson AP, Chang AR, Ciemins E, et al. Development and Validation of the American Heart Association’s PREVENT Equations. Circulation. 2024;149(6):430–449. doi:10.1161/CIRCULATIONAHA.123.067626

23. Samuels JM, Albaugh VL, Yu D, Chen Y, Williams DB, Spann MD, Wang L, Flynn CR, English WJ. Sex- and operation-dependent effects on 5-year weight loss results of bariatric surgery. Surgery for Obesity and Related Diseases. Published online January 2024:S1550728924000388. doi:10.1016/j.soard.2024.01.013

24. Docherty NG, Le Roux CW. Bariatric surgery for the treatment of chronic kidney disease in obesity and type 2 diabetes mellitus. Nat Rev Nephrol. 2020;16(12):709–720. doi:10.1038/s41581-020-0323-4

25. Goff DC, Lloyd-Jones DM, Bennett G, Coady S, D’Agostino RB, Gibbons R, Greenland P, Lackland DT, Levy D, O’Donnell CJ, et al. 2013 ACC/AHA Guideline on the Assessment of Cardiovascular Risk: A Report of the American College of Cardiology/American Heart Association Task Force on Practice Guidelines. Circulation. 2014;129(25_suppl_2). doi:10.1161/01.cir.0000437741.48606.98

26. Benotti PN, Wood GC, Carey DJ, Mehra VC, Mirshahi T, Lent MR, Petrick AT, Still C, Gerhard GS, Hirsch AG. Gastric Bypass Surgery Produces a Durable Reduction in Cardiovascular Disease Risk Factors and Reduces the Long-Term Risks of Congestive Heart Failure. JAHA. 2017;6(5):e005126. doi:10.1161/JAHA.116.005126

27. Persson CE, Björck L, Lagergren J, Lappas G, Giang KW, Rosengren A. Risk of Heart Failure in Obese Patients With and Without Bariatric Surgery in Sweden—A Registry-Based Study. Journal of Cardiac Failure. 2017;23(7):530–537. doi:10.1016/j.cardfail.2017.05.005

28. Sundström J, Bruze G, Ottosson J, Marcus C, Näslund I, Neovius M. Weight Loss and Heart Failure: A Nationwide Study of Gastric Bypass Surgery Versus Intensive Lifestyle Treatment. Circulation. 2017;135(17):1577–1585. doi:10.1161/CIRCULATIONAHA.116.025629

29. Jamaly S, Carlsson L, Peltonen M, Jacobson P, Karason K. Surgical obesity treatment and the risk of heart failure. European Heart Journal. 2019;40(26):2131–2138. doi:10.1093/eurheartj/ehz295

30. Berger S, Meyre P, Blum S, Aeschbacher S, Ruegg M, Briel M, Conen D. Bariatric surgery among patients with heart failure: a systematic review and meta-analysis. Open Heart. 2018;5(2):e000910. doi:10.1136/openhrt-2018-000910

31. Khan SS, Ning H, Wilkins JT, Allen N, Carnethon M, Berry JD, Sweis RN, Lloyd-Jones DM. Association of Body Mass Index With Lifetime Risk of Cardiovascular Disease and Compression of Morbidity. JAMA Cardiol. 2018;3(4):280. doi:10.1001/jamacardio.2018.0022

32. Masrur M, Bustos R, Sanchez-Johnsen L, Gonzalez-Ciccarelli L, Mangano A, Gonzalez-Heredia R, Patel R, Danielson KK, Gangemi A, Elli EF. Factors Associated with Weight Loss After Metabolic Surgery in a Multiethnic Sample of 1012 Patients. OBES SURG. 2020;30(3):975–981. doi:10.1007/s11695-019-04338-w

33. Hatoum IJ, Blackstone R, Hunter TD, Francis DM, Steinbuch M, Harris JL, Kaplan LM. Clinical Factors Associated With Remission of Obesity-Related Comorbidities After Bariatric Surgery. JAMA Surg. 2016;151(2):130. doi:10.1001/jamasurg.2015.3231

34. Hinerman AS, El Khoudary SR, Wahed AS, Courcoulas AP, Barinas-Mitchell EJM, King WC. Predictors of change in cardiovascular disease risk and events following gastric bypass: a 7-year prospective multicenter study. Surgery for Obesity and Related Diseases. 2021;17(5):910–918. doi:10.1016/j.soard.2020.12.013

35. Wolfe BM, Kvach E, Eckel RH. Treatment of Obesity: Weight Loss and Bariatric Surgery. Circulation Research. 2016;118(11):1844–1855. doi:10.1161/CIRCRESAHA.116.307591

36. Madsen LR, Baggesen LM, Richelsen B, Thomsen RW. Effect of Roux-en-Y gastric bypass surgery on diabetes remission and complications in individuals with type 2 diabetes: a Danish population-based matched cohort study. Diabetologia. 2019;62(4):611–620. doi:10.1007/s00125-019-4816-2

37. Still CD, Wood GC, Benotti P, Petrick AT, Gabrielsen J, Strodel WE, Ibele A, Seiler J, Irving BA, Celaya MP, et al. Preoperative prediction of type 2 diabetes remission after Roux-en-Y gastric bypass surgery: a retrospective cohort study. The Lancet Diabetes & Endocrinology. 2014;2(1):38–45. doi:10.1016/S2213-8587(13)70070-6

38. Echouffo-Tcheugui JB, Greene SJ, Papadimitriou L, Zannad F, Yancy CW, Gheorghiade M, Butler J. Population Risk Prediction Models for Incident Heart Failure: A Systematic Review. Circ: Heart Failure. 2015;8(3):438–447. doi:10.1161/CIRCHEARTFAILURE.114.001896

